# Event-related potential signatures of auditory predictive processing in first-episode psychosis: a passive pattern-based study

**DOI:** 10.64898/2026.10.19.26365874

**Authors:** Manuel Muñoz-Caracuel, Vanesa Muñoz, Brenda Angulo, Francisco J. Ruiz-Martinez, Salvador Perona-Garcelán, Marta I. Garrido, Rafael Romero-García, Antonio J. Vázquez Morejón, Benedicto Crespo-Facorro, Carlos M. Gómez

**Author notes:** Corresponding authors: Carlos M. Gómez., Benedicto Crespo-Facorro.

## Abstract

**Background:** Predictive-processing accounts propose altered prediction-error signaling in psychosis. However, event-related potential (ERP) findings in first-episode psychosis (FEP) are inconsistent and focused predominantly on mismatch negativity (MMN). Examining anticipatory and later post-stimulus ERP responses alongside MMN within a single passive paradigm may broaden the assessment of predictive processing in FEP.

**Methods:** 34 participants with non-affective FEP and 34 healthy controls (HC) underwent EEG during a passive auditory pattern-based paradigm. Mean amplitudes of contingent negative variation (CNV), MMN, and post-imperative negative variation (PINV) were quantified at five frontocentral electrodes. Exploratory analyses examined N1 and P2 mean amplitudes. Mixed models tested group, electrode, and group-by-electrode effects adjusting for age, sex and years of education. Regressions examined ERP associations with psychotic-like experiences, neurocognition, and social cognition across groups, and with symptoms, functioning, and antipsychotic dose within FEP.

**Results:** FEP participants reported more psychotic-like experiences and showed poorer global and social cognition than HC (all p-FDR ≤.042). Standardized adjusted FEP–HC differences at FCz were nonsignificant for CNV (β = −0.46), MMN (β = −0.30), and PINV (β = −0.10; all p-FDR ≥.265). Exploratory analyses identified lower P2 amplitude in FEP at FCz (β = −0.73; p-FDR =.019) and in the averaged frontocentral ROI (β = −0.64; p-FDR =.027). No ERP-phenotype associations survived FDR correction.

**Conclusions:** Despite phenotypic differences between FEP and HC, no significant differences emerged in the primary ERP indices of predictive processing examined. The exploratory P2 finding and its possible relationship to predictive processing and frontal dysfunction in FEP warrant further investigation.

## 1. INTRODUCTION

Predictive processing is an influential theoretical framework for understanding brain function ^1,2^. It proposes that the brain continuously generates and updates internal models of the world by integrating prior knowledge with incoming sensory information. Discrepancies between predicted and observed input are thought to be encoded as prediction errors that guide model updating, ultimately contributing to adaptive behavior ^3,4^. In psychosis research, this framework offers a computational account of how disturbances in the relative weighting of prior expectations and sensory evidence may underlie the emergence of hallucinations and delusions ^5–7^. However, whether neurophysiological responses associated with successive stages of predictive processing are altered in early psychosis remains unclear.

Auditory event-related potentials (ERPs) offer a temporally resolved approach to investigating neural processes relevant to predictive-processing accounts. Mismatch negativity (MMN), an ERP elicited by violations of auditory regularities even during passive listening ^8^, has been interpreted as a candidate index of prediction-error signaling within this framework ^9,10^. Robust reductions in MMN amplitude have been documented in established schizophrenia ^11^, generating interest in its potential as a neurophysiological biomarker ^12^. However, smaller and less consistent reductions in first-episode psychosis (FEP) and clinical high-risk populations raise questions about its sensitivity to early psychosis, while reductions in other psychiatric disorders raise concerns about its diagnostic specificity ^13^. Its interpretation as an index of prediction-error signaling is further complicated by physical differences between standard and deviant stimuli in conventional pitch-or duration-deviant oddball paradigms, which also allow MMN to be explained in terms of differential neural adaptation ^9^.

Pattern-based paradigms offer a complementary approach by defining deviance through violations of relationships among sounds, allowing physically identical tones to serve as standards or deviants depending on the preceding context. Such designs probe predictions based on sequential structure rather than solely on the adaptation to repetition of individual acoustic features ^14^. By requiring the extraction and integration of patterns across sounds, these paradigms may place greater demands on auditory processing, potentially revealing abnormalities in FEP that are less apparent in simpler paradigms ^15^. Consistent with this possibility, reduced MMN has been reported in early psychosis using different pattern-based paradigms ^16,17^, even in the presence of comparable pitch and duration deviant responses between patients and controls ^15^. Recent meta-analytic findings suggest that pattern-elicited MMN may be a more sensitive biomarker in the early phases of schizophrenia than the MMN elicited by conventional paradigms ^18^. However, the evidence remains limited by the small number of studies and heterogeneity of paradigms.

Importantly, even when complex auditory regularities are examined, the predominantly MMN-centered approach in the literature provides limited information about activity preceding and following deviance detection. Integrating MMN with anticipatory and slower post-stimulus responses may therefore offer a broader temporal perspective on predictive processing ^19^. In this context, contingent negative variation (CNV), associated with anticipation and preparation, has been investigated in schizophrenia, with meta-analytic evidence of reduced amplitude in paradigms involving attention and motor preparation ^20^. Post-imperative negative variation (PINV), associated with post-stimulus evaluation under uncertainty, has also shown alterations in schizophrenia during tasks involving stimulus ambiguity and uncertainty about response appropriateness ^21^. Together, these findings motivate extending ERP investigations in FEP beyond deviance detection. Whether comparable alterations occur during passive listening, and how they relate to predictive processing in early psychosis, remain unclear.

To address these questions, we used a passive auditory pattern-based oddball paradigm previously characterized by our group in healthy adults ^22^. A distinctive feature of this paradigm is that it elicits, during passive listening, a temporally ordered sequence of ERP responses compatible with successive stages of predictive processing: anticipation (CNV), context-dependent deviance detection (MMN), and subsequent slow post-stimulus updating (PINV). The present study extended this work to individuals with FEP and healthy controls. We examined group differences in frontocentral CNV, MMN, and PINV amplitudes and their associations with psychotic-like experiences, global neurocognition, and social cognition. Within the FEP group, we additionally examined relationships with symptom severity, social functioning, and current antipsychotic dose. Exploratory analyses extended these comparisons and associations to N1 and P2 ERP responses. By assessing these responses, we sought to determine whether candidate ERP indices of predictive processing are altered in FEP and whether they covary with clinically relevant dimensions.

## 2. METHODS

### 2.1. Participants

This cross-sectional case-control study included 68 participants: 34 individuals with first-episode non-affective psychosis (FEP) and 34 healthy comparison (HC) participants. Individuals with FEP were recruited from the Early Intervention Program for Psychosis at Hospital Universitario Virgen del Rocío, Seville, Spain. HC were matched to the FEP group by age and sex. Demographic and clinical characteristics are summarized in Table 1.

**Table 1.** Demographic, behavioral, and clinical characteristics of the study sample. Values are mean (SD) or n (%), as indicated. Longitudinal diagnoses reflect the diagnosis recorded at follow-up. CAPE-42, 42-item Community Assessment of Psychic Experiences; CPZ, chlorpromazine; FEP, first-episode psychosis; HC, healthy controls; MASC, Movie for the Assessment of Social Cognition; PANSS, Positive and Negative Syndrome Scale; SCIP, Screen for Cognitive Impairment

| Characteristic | HC (n = 34) | FEP (n = 34) |
| --- | --- | --- |
| **Sociodemographic variables** |  |  |
| Age, years | 24.41 (4.92) | 26.12 (6.02) |
| Male sex, n (%) | 25 (73.5) | 25 (73.5) |
| Education, years | 15.06 (2.03) | 12.88 (2.85) |
| **Primary behavioral measures** |  |  |
| CAPE-42 total score | 69.76 (12.43) | 78.76 (13.70) |
| SCIP total score | 86.94 (9.37) | 68.68 (12.14) |
| MASC correct score | 32.29 (3.69) | 26.09 (6.23) |
| **Clinical variables** |  |  |
| Time from FEP diagnosis, months | — | 7.47 (6.52) |
| Current antipsychotic treatment: yes, n (%) | — | 31 (91.2) |
| Antipsychotic dose, CPZ equivalents, mg/day | — | 220.41 (166.27) |
| PANSS positive scale | — | 10.29 (3.18) |
| PANSS negative scale | — | 12.56 (4.03) |
| PANSS general psychopathology | — | 26.06 (4.28) |
| SFS total score | — | 110.62 (16.50) |
| **Longitudinal diagnosis, n (%)** |  |  |
| Brief psychotic disorder | — | 14 (41.2) |
| Schizophrenia | — | 13 (38.2) |
| Unspecified psychotic disorder | — | 3 (8.8) |
| Schizophreniform disorder | — | 2 (5.9) |
| Delusional disorder | — | 1 (2.9) |
| Schizotypal personality disorder | — | 1 (2.9) |

Eligibility criteria for the FEP group were: (1) age between 18 and 36 years; (2) meeting DSM-5 criteria for a schizophrenia spectrum or other psychotic disorder, including schizophrenia, schizophreniform disorder, brief psychotic disorder, delusional disorder, schizotypal personality disorder, or unspecified psychotic disorder; (3) first diagnosis of psychosis within the preceding 24 months; and (4) no more than 12 cumulative months of antipsychotic treatment before study enrollment. Diagnoses were confirmed using the Structured Clinical Interview for DSM-5 (SCID-5), administered by trained psychiatrists or clinical psychologists. The initial diagnosis was reassessed 6 months after enrollment and revised during follow-up when clinically indicated. Antipsychotic medication was prescribed at the minimum effective dose according to routine clinical practice. The diagnostic distribution of the analyzed sample and current antipsychotic doses at the time of EEG assessment are reported in Table 1.

Exclusion criteria for all participants were: (1) a current DSM-5 substance use disorder other than tobacco and cannabis use disorder; (2) a history of a neurological disorder; (3) self-reported hearing impairment; and (4) affective first-episode psychosis. Healthy comparison participants were additionally required to have no current or lifetime psychiatric disorder. The study was approved by the Research Ethics Committee of the Junta de Andalucía (reference 1045-N-21) and was conducted in accordance with the Declaration of Helsinki. All participants provided written informed consent before enrollment.

### 2.2. Clinical, neurocognitive and social-cognitive measures

The cross-group phenotypic assessment included instruments suitable for both clinical and nonclinical populations to characterize psychotic-like experiences, neurocognitive performance, and social cognition dimensionally. Validated Spanish-language versions were used: the 42-item Community Assessment of Psychic Experiences (CAPE-42), measuring self-reported psychotic-like experiences ^23^; the Screen for Cognitive Impairment in Psychiatry (SCIP-S), assessing global neurocognitive performance ^24^; and the Movie for the Assessment of Social Cognition (MASC), assessing social cognition ^25^. Additional assessments within the FEP group included symptom severity, evaluated using the Positive and Negative Syndrome Scale (PANSS) ^26^ by trained psychiatrists or clinical psychologists, and social functioning, measured using the self-report version of the Social Functioning Scale (SFS) ^27^. Antipsychotic doses at the time of EEG assessment were converted into chlorpromazine-equivalent daily doses ^28^.

### 2.3. EEG

#### 2.3.1. Auditory predictive-processing paradigm

We used a passive auditory pattern-based oddball paradigm previously characterized by our group in healthy adults ^22^. This paradigm has been shown to elicit a temporally ordered sequence of ERPs compatible with successive stages of predictive-processing: anticipation (CNV), context-dependent deviance detection (MMN), and subsequent post-stimulus updating (PINV) ^19^.

The paradigm was programmed using PsychoPy (version 2021.1.2), and auditory stimuli were prepared using Audacity (version 3.0.0). Each trial comprised four consecutive pure tones (S1–S4) arranged in an ascending (600, 900, 1200, and 1500 Hz) or descending pattern (1500, 1200, 900, and 600 Hz), randomly counterbalanced. Each tone lasted 200 ms, including 20-ms rise and fall times, and tones were separated by a 100-ms interstimulus interval. Consequently, S2, S3, and S4 occurred 300, 600, and 900 ms after S1 onset, respectively. The interval between S4 offset and the next trial onset was 2900 ms, yielding a 4000-ms interval between consecutive trial onsets.

In standard trials, S4 conformed to the ascending or descending pattern established by S1–S3, whereas in deviant trials it violated this pattern (S4 = S1; Figure 1A). In addition to uncertainty about S4 type (standard or deviant), uncertainty about sequence direction (ascending or descending) required auditory predictions to be established on a trial-by-trial basis. Crucially, physically identical S4 tones could serve as standards or deviants depending on the preceding S1–S3 context. This design therefore constrained explanations of mismatch activity based on neural adaptation or habituation, supporting its interpretation in terms of context-dependent prediction-error signaling.

**Figure 1.**
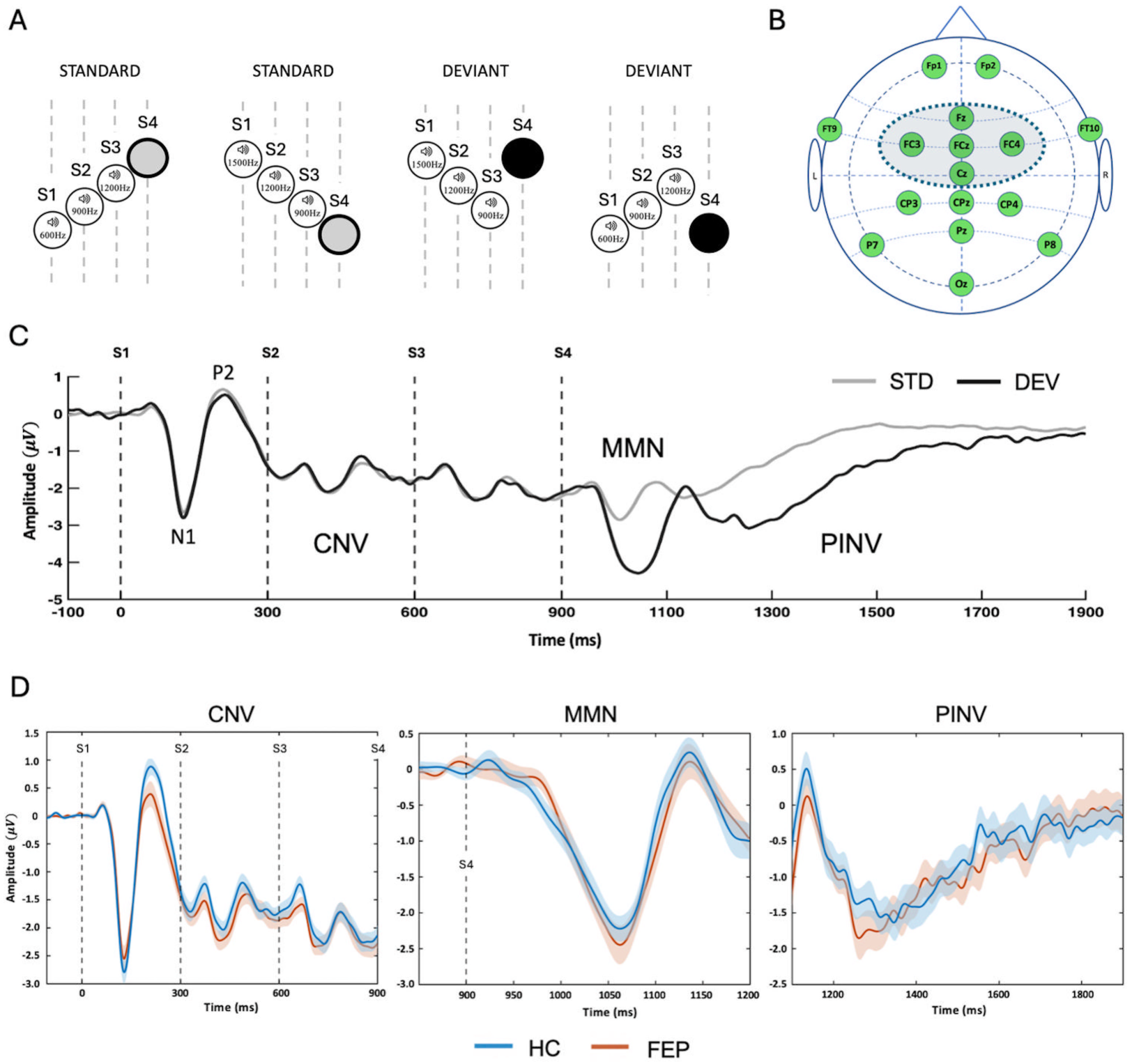
(A) Standard and deviant four-tone sequences (S1–S4) included in the experimental paradigm, with S4 status determined by the preceding S1-S3 context. (B) Electrode montage highlighting the five frontocentral sites analyzed. (C) Grand-average standard (gray) and deviant (black) waveforms at FCz, collapsed across groups, showing the temporal sequence of the ERP components examined. (D) Waveforms at FCz for healthy controls (blue) and first-episode psychosis participants (orange). CNV was averaged across trial types; MMN and PINV are deviant-minus-standard waveforms. Baselines were 100 ms pre-S1 for CNV, 50 ms pre-S4 for MMN, and 100 ms pre-S4 for PINV. All times are relative to S1 onset. CNV, contingent negative variation; MMN, mismatch negativity; PINV, post-imperative negative variation.

Each participant completed 400 trials: 350 standard trials (87.5%) and 50 deviant trials (12.5%). Trial order was pseudorandomized, with six to eight consecutive standard trials between successive deviant trials. Participants were instructed to ignore the sounds and attend to a muted film throughout the task. Auditory stimuli were presented at 60 dB SPL through two loudspeakers positioned on either side of the participant and calibrated at the listening position using a sound-level meter.

#### 2.3.2 Signal acquisition and preprocessing

EEG recordings and phenotypic assessments in participants with FEP were conducted during the post-acute phase, following sufficient clinical stabilization. EEG was recorded from 16 active scalp electrodes using an ActiCAP system (Figure 1B). Signals were amplified using a V-Amp DC amplifier (Brain Products GmbH, Munich, Germany), referenced online to the left mastoid, and sampled at 1000Hz with a gain of 20,000. Electrode impedances were maintained below 20 kΩ. Data were acquired using BrainVision Recorder 1.20 (Brain Products GmbH).

EEG preprocessing was performed in EEGLAB (version 2026.0.0) and MATLAB R2025b. Continuous data were visually inspected, and segments containing marked signal discontinuities or saturation were manually removed. Persistently noisy channels were interpolated using spherical spline interpolation. Continuous data were subsequently re-referenced offline to the common average and band-pass filtered from 0.05 to 30 Hz using a zero-phase FIR filter. Independent component analysis (ICA) was then applied to continuous data. Components were classified using ICLabel, and those labelled as eye or muscle with probability ≥.80 were removed automatically. Details of the removed ICA components, manually excluded segments, and interpolated channels are provided in Supplementary Data, Table S1.

Following ICA correction, two parallel epoching and artifact-rejection branches were created from the same continuous EEG data to accommodate the different temporal windows required for MMN and the slower CNV and PINV responses. The short branch, used for the MMN analyses, comprised epochs from −50 to 300 ms relative to S4. The long branch, used for the CNV and PINV analyses, extended from −100 ms before S1 to 1000 ms after S4. Epochs exceeding ±100 µV at any of the 16 channels were automatically rejected. Participant-level artifact-rejection metrics are reported in Supplementary Data, Table S2.

#### 2.3.3. ERP quantification

ERP measures were quantified at five frontocentral electrodes (Fz, FCz, Cz, FC3, and FC4; Figure 1B), selected on the basis of the well-established frontocentral distribution of the components under investigation (CNV, MMN, and PINV) ^8,21,29^, and evidence of maximal amplitudes at frontocentral sites in similar experimental paradigms ^22,30,31^. For the primary ERP analyses, CNV was quantified as the mean amplitude from S1-S4 (0-900 ms), averaged across standard and deviant trials. MMN was quantified as the mean deviant-minus-standard amplitude within a 100-ms window (112-212 ms after S4) centered on the negative peak identified at 162 ms in the FCz grand-average difference waveform collapsed across groups and conditions. PINV was quantified as the mean deviant-minus-standard amplitude from 300 to 1000 ms after S4. Additional exploratory analyses examined N1 and P2 mean amplitudes after S1 within separate 50-ms windows centered on their respective peaks (120 and 195 ms after S1, respectively) in the FCz waveform collapsed across groups and conditions. Peak amplitudes, latencies, and mean amplitudes within shorter windows for CNV (the 100 ms preceding S4) and PINV (300-500 ms after S4) were also explored.

#### 2.3.4. Statistical Analysis

Statistical analyses were performed using R software (version 4.6.1). Cross-group differences in the primary clinical, neurocognitive, and social-cognitive measures were estimated using linear models adjusted for age, sex, and years of education. For each primary ERP measure (CNV, MMN, and PINV), electrode-level amplitudes were analyzed using linear mixed-effects models including group, electrode, the group-by-electrode interaction, age, sex, and years of education as fixed effects and a participant-specific random intercept. The Benjamini-Hochberg procedure ^32^ was used to control the false discovery rate (FDR) separately within three prespecified families of tests: (1) the three primary clinical, neurocognitive, and social-cognitive group comparisons; (2) the electrode-specific group contrasts for the primary ERP measures (three ERP measures × five electrodes); and (3) the three group-by-electrode interaction tests, one for each primary ERP measure. Additional clinical subscales and exploratory ERP measures were analyzed and corrected within their respective analytical families. When a group-by-electrode interaction survived FDR correction, pairwise post hoc comparisons between electrode-specific FEP-HC contrasts were adjusted using the Holm procedure.

For the EEG-phenotype analyses, pooled linear models regressed each outcome on ERP amplitude, group, age, sex, and years of education, thereby estimating a common ERP-outcome association across groups. Separate models included the ERP-by-group interaction to examine whether these associations differed between FEP and HC participants. FEP-only models also examined ERP associations with symptom severity and social functioning. Within the FEP group, separate models examined associations between current chlorpromazine-equivalent dose and ERP and phenotypic measures. Additionally, ERP-outcome models within FEP included dose as a covariate to assess whether this adjustment altered the association estimates.

## 3. Results

### 3.1 Clinical, neurocognitive and social-cognitive measures

Descriptive statistics are presented in Table 1. After adjustment for age, sex, and education, participants with FEP reported higher CAPE-42 total scores than HC (standardized adjusted group difference, β = 0.59; 95% CI, 0.02 to 1.16; p-FDR =.042). Participants with FEP also showed poorer global neurocognitive performance, indexed by lower SCIP total scores (β = −1.22; 95% CI, −1.71 to −0.72; p-FDR <.001), and poorer social-cognitive performance, reflected in lower MASC correct scores (β = −0.72; 95% CI, −1.23 to −0.22; p-FDR =.009) (Figure 2A). Additional results are reported in Supplementary Data, Table S3.

**Figure 2.**
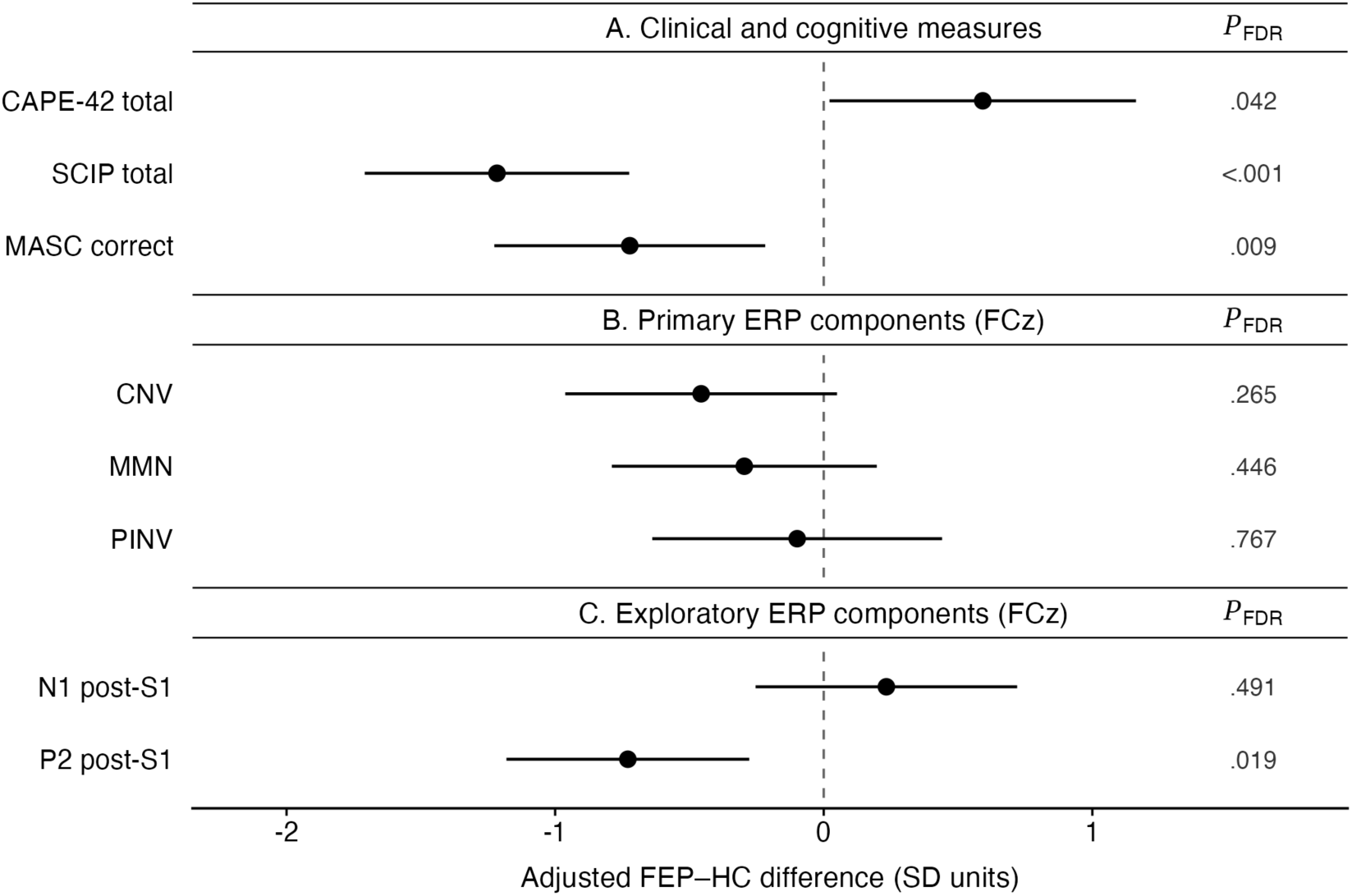
Adjusted FEP-HC differences in clinical and cognitive measures (A), primary ERP mean amplitudes at FCz (B), and exploratory N1 and P2 mean amplitudes following S1 at FCz (C). Points and lines indicate adjusted differences and 95% confidence intervals in pooled standard-deviation units, estimated using linear models (A) and mixed-effects models (B and C), adjusted for age, sex, and education. FDR correction was applied separately to three clinical/cognitive comparisons, 15 primary ERP comparisons, and 10 exploratory N1/P2 comparisons. Confidence intervals are not adjusted for multiple comparisons. FEP, first-episode psychosis; HC, healthy controls; FDR, false discovery rate.

### 3.2. ERPs

Grand-average waveforms at FCz collapsed across FEP and HC participants showed he expected ERP activity, including N1 and P2 responses following S1, a sustained negative potential from S1-S4 compatible with CNV, an early deviance-related negativity following S4 compatible with MMN, and a subsequent sustained negativity compatible with PINV (Figure 1C). Group-specific waveforms illustrating the corresponding responses in FEP and HC participants at FCz are presented in Figure 1D. Corresponding waveforms at each of the five frontocentral electrodes and averaged across the frontocentral ROI are shown in Supplementary Material, Figures S1-S3 for CNV, MMN, and PINV, respectively. Responses to standard and deviant S4 tones in FEP and HC participants are shown in Figures S4 and S5.

At FCz, age, sex, and education adjusted FEP–HC differences were not significant for CNV mean amplitude (standardized adjusted difference, β = −0.46; 95% CI, −0.96 to 0.05; p-FDR =.265), MMN mean amplitude (β = −0.30; 95% CI, −0.79 to 0.20; p-FDR =.446), or PINV mean amplitude (β = −0.10; 95% CI, −0.64 to 0.44; p-FDR =.767) (Figure 2B). Likewise, no group differences in these primary measures survived FDR correction at the other electrodes of interest (Fz, Cz, FC3, and FC4) or in the averaged frontocentral ROI. No group-by-electrode interaction survived FDR correction for the primary ERP measures (Supplementary Data, Table S3).

Exploratory analyses of N1 and P2 identified lower P2 mean amplitude following S1 in FEP than HC at FCz (β = −0.73; 95% CI, −1.18 to −0.28; p-FDR =.019; Figure 2C) and in the averaged frontocentral ROI (β = −0.64; 95% CI, −1.14 to −0.13; p-FDR =.027). No significant group differences were observed for N1 mean amplitude. An exploratory group-by-electrode interaction was observed for MMN peak amplitude, F(4, 264) = 4.51, p-FDR =.009, indicating that the adjusted FEP-HC difference varied across electrodes. Holm-adjusted post hoc comparisons revealed more negative FEP–HC contrasts at Fz (p <.001), FCz (p =.043), Cz (p =.043), and FC3 (p =.034) than at FC4, although no electrode-specific group contrast survived correction. Full exploratory results and post hoc comparisons are reported in Supplementary Data, Table S4.

### 3.3. Behavioral and electrophysiological associations

After FDR correction, primary ERP mean amplitudes at FCz were not significantly associated with psychotic-like experiences, global neurocognitive performance, or social cognition in pooled models adjusted for group, age, sex, and education, or with symptom severity or social functioning in FEP-only models adjusted for age, sex and education. The same pattern was observed for both total scores and individual subscales (Table 2). No significant associations emerged at the remaining frontocentral electrodes or the averaged frontocentral ROI after correction for multiple comparisons. Exploratory analyses of N1 and P2 mean amplitudes following S1 likewise revealed no associations with clinical, neurocognitive, social-cognitive, or functional measures that survived FDR correction (Supplementary Data, Table S5).

**Table 2.** Associations of ERP mean amplitudes at FCz electrode with clinical, neurocognitive, social-cognitive and functional outcomes. β values are standardized regression coefficients. CAPE-42, SCIP, and MASC models included both groups and were adjusted for group, age, sex, and education. SFS and PANSS models included the FEP group only and were adjusted for age, sex, and education. Benjamini–Hochberg FDR correction was applied separately within each prespecified analytical family, comprising associations of the three ERP measures with either the primary total scores or each instrument’s subscales. CNV, contingent negative variation; MMN, mismatch negativity; PINV, post-imperative negative variation; CI, confidence interval; FDR, false discovery rate.

| Outcome | CNV |  |  | MMN |  |  | PINV |  |  |
| --- | --- | --- | --- | --- | --- | --- | --- | --- | --- |
| | $\beta$ (95% CI) | p | p-FDR | $\beta$ (95% CI) | p | p-FDR | $\beta$ (95% CI) | p | p-FDR |
| **CAPE-42 total score** | 0.08 (−0.17, 0.33) | .537 | .887 | 0.02 (−0.23, 0.26) | .887 | .887 | 0.02 (−0.23, 0.27) | .880 | .887 |
| Positive subscale | 0.12 (−0.14, 0.37) | .367 | .701 | −0.17 (−0.41, 0.08) | .184 | .701 | −0.09 (−0.34, 0.17) | .512 | .701 |
| Negative subscale | 0.02 (−0.24, 0.27) | .881 | .881 | 0.07 (−0.17, 0.32) | .544 | .701 | 0.05 (−0.20, 0.31) | .680 | .765 |
| Depressive subscale | 0.08 (−0.18, 0.33) | .545 | .701 | 0.18 (−0.06, 0.42) | .140 | .701 | 0.10 (−0.15, 0.36) | .415 | .701 |
| **SCIP total score** | 0.07 (−0.10, 0.25) | .418 | .887 | 0.02 (−0.15, 0.19) | .824 | .887 | −0.03 (−0.21, 0.14) | .722 | .887 |
| Verbal learning—immediate | 0.20 (0.01, 0.40) | .044 | .658 | 0.03 (−0.17, 0.23) | .754 | .918 | 0.05 (−0.16, 0.25) | .644 | .918 |
| Working memory | −0.03 (−0.27, 0.20) | .769 | .918 | −0.03 (−0.25, 0.19) | .790 | .918 | −0.13 (−0.36, 0.10) | .280 | .918 |
| Verbal fluency | 0.03 (−0.19, 0.25) | .786 | .918 | −0.02 (−0.23, 0.19) | .857 | .918 | −0.04 (−0.27, 0.18) | .688 | .918 |
| Verbal learning—delayed | 0.06 (−0.16, 0.29) | .584 | .918 | 0.13 (−0.08, 0.35) | .224 | .918 | 0.08 (−0.15, 0.30) | .489 | .918 |
| Processing speed | 0.00 (−0.18, 0.18) | .998 | .998 | 0.02 (−0.16, 0.19) | .837 | .918 | −0.05 (−0.23, 0.13) | .586 | .918 |
| **MASC correct score** | 0.04 (−0.17, 0.24) | .725 | .887 | 0.14 (−0.05, 0.33) | .139 | .887 | 0.02 (−0.18, 0.23) | .811 | .887 |
| No-mentalizing errors | −0.01 (−0.20, 0.18) | .938 | .974 | 0.00 (−0.18, 0.19) | .974 | .974 | 0.03 (−0.16, 0.22) | .733 | .943 |
| Undermentalizing errors | 0.05 (−0.18, 0.29) | .645 | .943 | −0.21 (−0.43, 0.01) | .062 | .559 | −0.16 (−0.39, 0.08) | .185 | .833 |
| Overmentalizing errors | −0.10 (−0.32, 0.11) | .347 | .943 | −0.07 (−0.28, 0.14) | .500 | .943 | 0.06 (−0.15, 0.28) | .566 | .943 |
| **SFS total** | 0.12 (−0.31, 0.56) | .567 | .920 | 0.02 (−0.37, 0.41) | .920 | .920 | 0.07 (−0.32, 0.46) | .724 | .920 |
| Social engagement/withdrawal | 0.32 (−0.09, 0.72) | .120 | .733 | 0.09 (−0.29, 0.46) | .632 | .948 | 0.25 (−0.12, 0.61) | .174 | .733 |
| Interpersonal communication | 0.10 (−0.29, 0.50) | .604 | .948 | −0.03 (−0.39, 0.32) | .851 | .948 | 0.06 (−0.29, 0.42) | .713 | .948 |
| Independence—performance | −0.15 (−0.55, 0.26) | .462 | .883 | 0.20 (−0.15, 0.56) | .252 | .733 | 0.03 (−0.33, 0.39) | .867 | .948 |
| Independence—competence | 0.03 (−0.36, 0.42) | .886 | .948 | 0.28 (−0.06, 0.61) | .099 | .733 | 0.20 (−0.14, 0.54) | .243 | .733 |
| Recreation | 0.22 (−0.21, 0.64) | .314 | .733 | 0.07 (−0.33, 0.46) | .733 | .948 | 0.23 (−0.15, 0.61) | .234 | .733 |
| Prosocial activities | 0.10 (−0.31, 0.52) | .608 | .948 | −0.33 (−0.68, 0.02) | .067 | .733 | −0.19 (−0.55, 0.18) | .300 | .733 |
| Employment/occupation | −0.02 (−0.42, 0.37) | .903 | .948 | 0.00 (−0.35, 0.36) | .982 | .982 | −0.14 (−0.49, 0.21) | .412 | .865 |
| **PANSS positive scale** | 0.15 (−0.25, 0.55) | .450 | .860 | −0.23 (−0.58, 0.13) | .200 | .860 | −0.10 (−0.46, 0.26) | .578 | .860 |
| **PANSS negative scale** | −0.06 (−0.43, 0.32) | .764 | .860 | −0.14 (−0.47, 0.19) | .395 | .860 | −0.02 (−0.35, 0.32) | .920 | .920 |
| **PANSS general psychopathology** | 0.13 (−0.28, 0.53) | .518 | .860 | −0.11 (−0.48, 0.25) | .525 | .860 | 0.06 (−0.30, 0.43) | .728 | .860 |

### 3.4. Medication-related analyses

Current chlorpromazine-equivalent dose was not significantly associated with the primary ERP measures or exploratory N1 and P2 mean amplitudes after adjustment for age, sex, and education and FDR correction. Additional adjustment for dose did not change the conclusions regarding ERP-outcome associations within the FEP group (Supplementary Data, Table S6).

## 4. Discussion

This study examined candidate ERP indices of auditory predictive processing in FEP by assessing anticipatory, deviance-related, and slower post-stimulus responses within a single passive pattern-based paradigm. Despite reporting more psychotic-like experiences and showing poorer neurocognitive and social-cognitive performance, participants with FEP showed no significant differences from HC in the primary CNV, MMN, or PINV mean amplitudes after adjustment for age, sex and education. These findings were consistent across the five frontocentral electrodes analyzed and the averaged frontocentral ROI. In contrast, exploratory analyses identified lower P2 mean amplitude in FEP at FCz and in the averaged frontocentral ROI. No ERP-phenotype associations survived FDR correction.

Emerging evidence of MMN reductions with pattern-based paradigms has generated interest in their potential to detect auditory processing abnormalities in early psychosis ^18^. This is particularly relevant because MMN reductions elicited by conventional paradigms in FEP have been smaller and less consistent than those observed in established schizophrenia ^33,34^. However, we found no significant MMN amplitude differences between FEP and HC. Similarly, Salisbury et al. ^35^ found reduced MMN to violations of ascending tone-pair relationships in long-term schizophrenia but no significant reduction in first-episode schizophrenia-spectrum psychosis. Collectively, these observations suggest that pattern-based MMN reductions are not consistently observed at first episode and that their detection may depend on specific paradigm characteristics.

Several differences warrant consideration when comparing these findings with previous reports. Our paradigm examined violations of a four-tone frequency trajectory rather than omitted or additional tones used in previous studies. The randomized presentation of ascending and descending sequences in our study also introduced uncertainty about sequence direction at trial onset, requiring predictions to be established from the unfolding context of each trial. These features may engage auditory processing differently from paradigms based on a stable pattern repeated across trials. Component definition is another source of heterogeneity. Some pattern-based studies have reported reductions in a later deviance-related negativity, described as late complex MMN, approximately 350–450 ms after the violation, without significant reductions in the earlier MMN response ^36,37^. These later responses overlap temporally with the early PINV window examined here (300–500 ms after S4), highlighting differences in component definition that complicate direct comparisons. However, neither MMN nor early PINV showed significant group differences after FDR correction in our study. Clinical differences further constrain comparisons: some studies included participants within the first five years of illness ^16,17^, whereas our sample was restricted to individuals within two years of the first clinical identification of psychosis, with a mean interval of approximately seven months. Variation in diagnostic composition and clinical characteristics may further contribute to differences in MMN findings across early-psychosis samples.

A distinctive contribution of this study was the assessment of slower activity preceding and following deviance detection within a single passive auditory paradigm, without requiring a task-related behavioral response. Within a predictive-processing framework, CNV and PINV have been interpreted as candidate indices of anticipation and post-stimulus evaluation or updating, respectively, extending the temporal assessment beyond MMN ^19^. In the present study, neither CNV nor PINV mean amplitude differed significantly between groups after FDR correction. Although CNV attenuation and PINV alterations have been reported in schizophrenia during active tasks involving attention ^20,21^, their generalizability to passive auditory processing in FEP remains uncertain. The absence of significant CNV differences in our sample parallels the findings of Kirenskaya et al. ^38^, who observed significant frontal CNV reductions during an active task in chronic but not first-episode schizophrenia. By contrast, Li et al. ^39^ reported reduced CNV amplitude and altered PINV measures in first-episode, drug-naïve patients with deficit schizophrenia, a more narrowly defined clinical subgroup than the broader FEP sample examined here.

The computational interpretation of these findings requires caution. Although the paradigm captures temporally distinct ERP responses, interpreting CNV, MMN, and PINV as candidate indices of anticipation, prediction-error signaling, and subsequent updating does not establish a direct correspondence between their amplitudes and specific computational quantities ^19^. The present analyses neither separately estimated the precision of prior expectations and sensory evidence nor quantified trial-by-trial amplitude changes. Different combinations of changes in prior and sensory precision may produce similar patterns of information weighting ^7^, which amplitude comparisons alone cannot disentangle. Consequently, the absence of significant group differences in CNV, MMN, and PINV should not be interpreted as evidence that predictive processing is intact, but as indicating that these primary measures did not detect group-level alterations under the conditions studied.

The exploratory P2 findings warrant particular attention. Participants with FEP showed lower P2 mean amplitude following S1 both at FCz and in the averaged frontocentral ROI. The direction of this difference is consistent with reductions in P2 amplitude reported in first-episode schizophrenia ^40^ and with meta-analytic findings in schizophrenia ^41^. P2 modulation has also been associated with auditory stimulus history and contextual regularity, suggesting a potential role in the establishment of auditory expectations ^42,43^. Additional evidence points to a possible prefrontal contribution to this component, supported by neurovascular coupling between P2 and prefrontal hemodynamic responses assessed using simultaneous EEG and functional near-infrared spectroscopy (fNIRS) ^44,45^. This possibility is relevant because impaired prefrontal recruitment during contextual processing has been documented in first-episode schizophrenia ^46^, and disrupted prefrontal connectivity has been associated with cognitive impairment, disorganization, and poorer global functioning ^47^. A passive electrophysiological measure sensitive to such dysfunction could complement task-based assessments while reducing dependence on task comprehension and performance. In our paradigm, the randomized presentation of ascending and descending sequences made S1 informative about sequence direction, providing a basis for establishing predictions about subsequent tones within each trial. The lower P2 amplitude in FEP may therefore reflect differences in the initial processing of this predictive context. However, no P2-phenotype associations survived FDR correction, leaving its clinical relevance uncertain. Future studies combining manipulations of the predictive information conveyed by S1 with concurrent measures of prefrontal activity could clarify whether P2 reflects frontal contributions to auditory expectation formation and whether this relationship has clinical relevance in FEP.

A separate exploratory group-by-electrode interaction for MMN peak amplitude indicated variation in the spatial pattern of group differences, although no electrode-specific contrast survived correction. This interaction should be interpreted cautiously, as the corresponding interaction for mean amplitude did not survive FDR correction and peak estimates are particularly sensitive to waveform noise ^48^.

The phenotypic analyses provided a complementary dimensional assessment. Pooled models adjusted for group, age, sex, and education assessed whether individual variation in ERP amplitudes was associated with phenotypic variation beyond average differences between FEP and HC, assuming a common slope across groups. Despite substantial group differences in psychotic-like experiences, neurocognition, and social cognition, no associations with the primary or exploratory ERP measures survived FDR correction. Within FEP, no associations with symptom severity or social functioning survived FDR correction. Overall, these results do not establish the ERP measures examined as dimensional markers of the assessed phenotypic characteristics.

Current chlorpromazine-equivalent dose was not significantly associated with the primary ERP measures or exploratory N1 and P2 mean amplitudes, and additional adjustment for dose did not materially alter the ERP-outcome associations within FEP. However, these findings do not completely exclude medication effects. Relying on current dose alone may overlook cumulative exposure, treatment duration, adherence, or pharmacological differences between compounds, and adjusting for current dose cannot fully disentangle medication effects from the clinical factors influencing prescribing.

Several limitations constrain interpretation. First, the sample of 34 participants per group may have provided insufficient statistical power to detect small-to-moderate group differences or ERP-phenotype associations, particularly in FEP-only analyses and after correction for multiple comparisons. Second, the cross-sectional design precludes determining whether ERP characteristics change with illness progression or predict subsequent outcomes, while the lack of a chronic schizophrenia comparison group limits comparisons across illness stages. Third, the absence of a conventional oddball comparison condition prevents direct assessment of the relative sensitivity of the present pattern-based paradigm. Larger longitudinal studies spanning illness stages and combining independent manipulations of contextual predictability and sensory uncertainty with computational modeling could clarify how ERP responses relate to information weighting and model updating within the predictive-processing framework.

### 4.1. Conclusion

Despite clinical, neurocognitive, and social-cognitive differences between FEP and HC, this study found no robust evidence of group differences in the primary ERP indices of predictive processing examined (CNV, MMN, and PINV) under the passive pattern-based paradigm used. No ERP-phenotype associations survived FDR correction. Exploratory analyses identified lower P2 amplitude in FEP, although its possible relationship to predictive processing and frontal dysfunction requires further investigation.

## Data Availability

All data produced in the present study are available upon reasonable request to the authors

https://github.com/mmunozcaracuel/ERPs-auditory-predictive-processing-FEP

## ACKNOWLEDGEMENTS

The authors thank all the participants for their time, patience, and contribution to this study.

## DATA AND CODE AVAILABILITY

All code used to perform the analyses can be found at https://github.com/mmunozcaracuel/ERPs-auditory-predictive-processing-FEP. Data are available upon request.

## AUTHOR CONTRIBUTIONS

MM-C and CMG conceived and designed the experiment. MM-C, VM, BA and FJR-M conducted the experiment. MM-C, SP-G, AJVM, and BC-F recruited FEP participants. MM-C, VM, and BA recruited HC participants. MM-C analyzed the data. RR-G provided statistical and methodological support. MIG contributed to the theoretical and conceptual framework. MM-C drafted the manuscript under the supervision of CMG and BC-F. All authors revised the manuscript, approved the final version, and agree to be accountable for all aspects of the work, ensuring that questions related to the accuracy or integrity of any part of the work are appropriately investigated and resolved. All persons designated as authors qualify for authorship, and all those who qualify for authorship are listed.

