## Supplementary Material for "Event-related potential signatures of auditory predictive processing in first-episode psychosis: a passive pattern-based study"

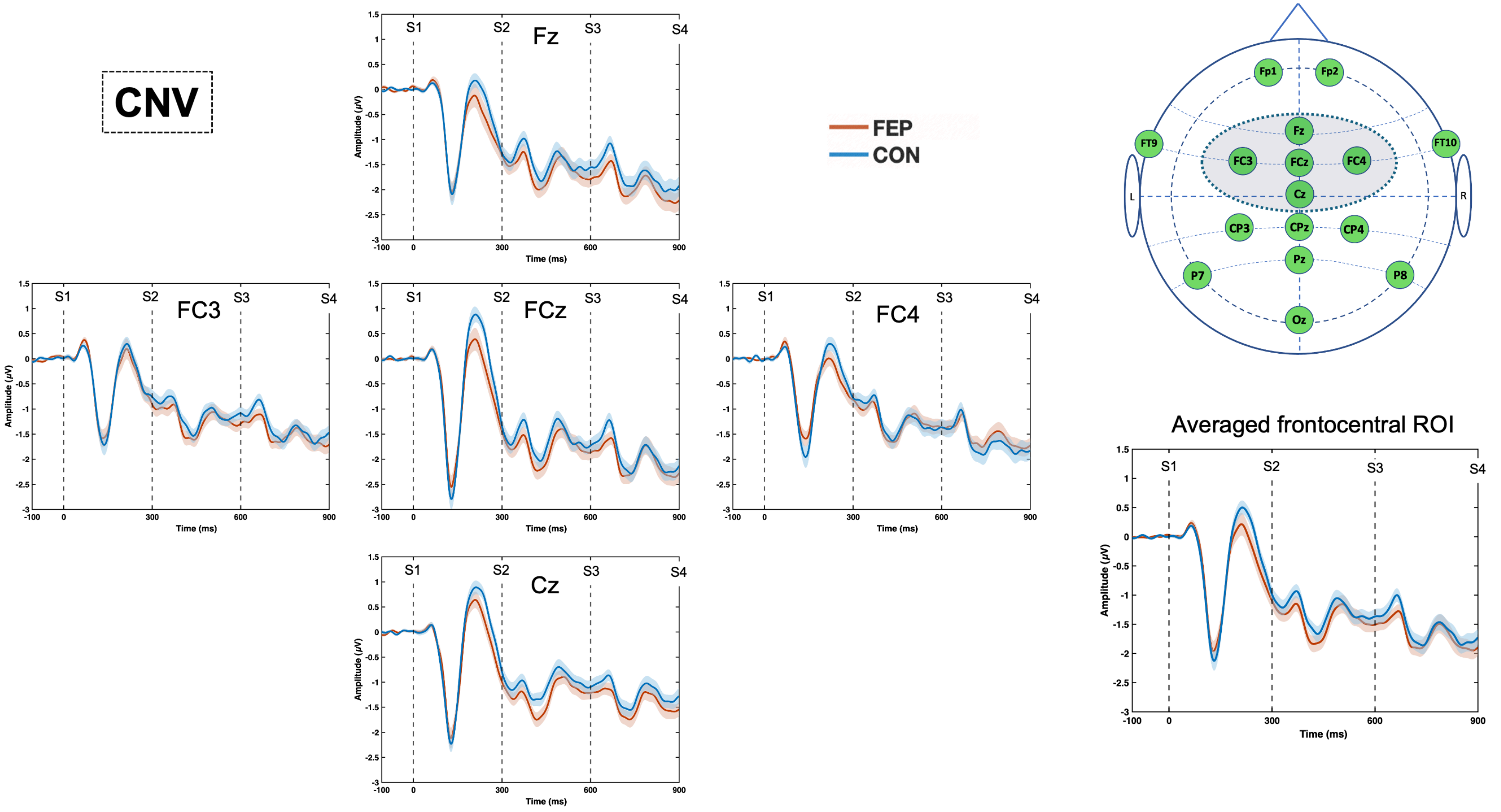

**Figure S1.** Grand-average waveforms illustrating contingent negative variation (CNV), averaged across standard and deviant trials, at five frontocentral electrodes (Fz, FCz, Cz, FC3, and FC4) and across the frontocentral region of interest (ROI). Lines represent group means, and shaded bands indicate ±1 SEM. The scalp diagram identifies the electrodes included in the ROI. FEP, first-episode psychosis; HC, healthy controls.

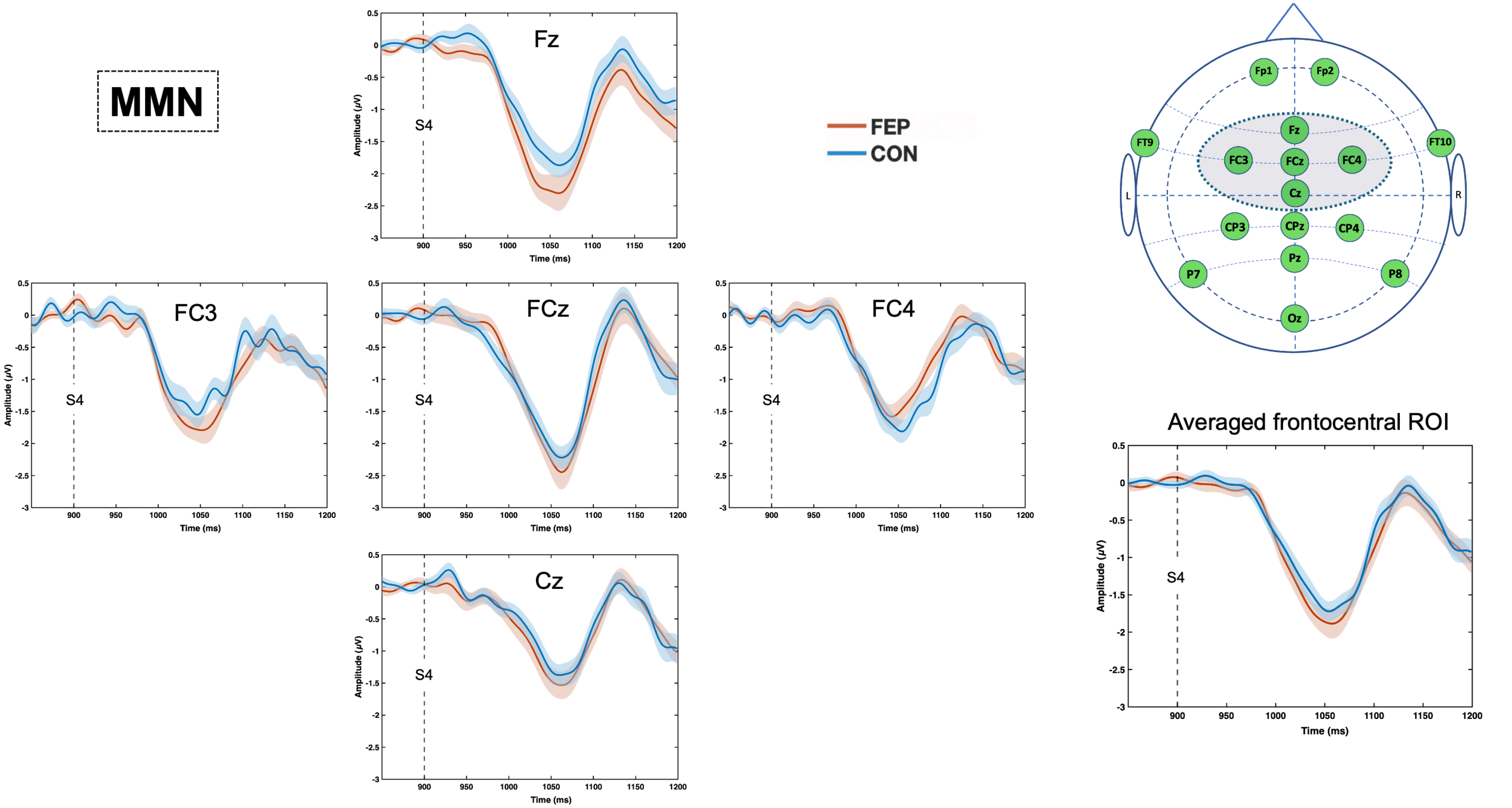

**Figure S2.** Grand-average deviant-minus-standard difference waveforms illustrating mismatch negativity (MMN) at five frontocentral electrodes (Fz, FCz, Cz, FC3, and FC4) and across the frontocentral region of interest (ROI). Lines represent group means, and shaded bands indicate ±1 SEM. The vertical dashed line marks S4 onset. The scalp diagram identifies the electrodes included in the ROI. FEP, first-episode psychosis; HC, healthy controls.

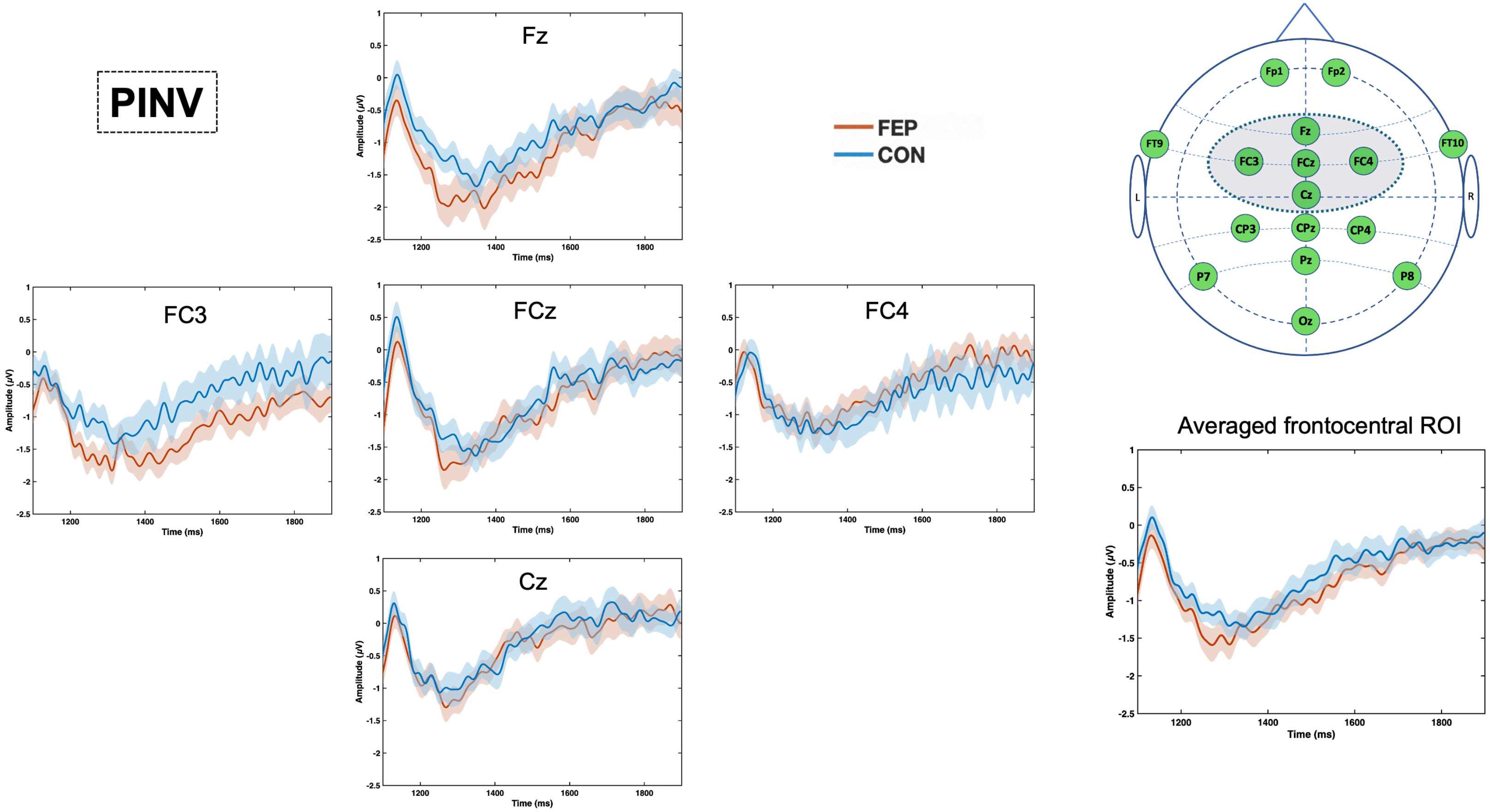

**Figure S3.** Grand-average deviant-minus-standard difference waveforms illustrating post-imperative negative variation (PINV) at five frontocentral electrodes (Fz, FCz, Cz, FC3, and FC4) and across the frontocentral region of interest (ROI). Lines represent group means, and shaded bands indicate ±1 SEM. The scalp diagram identifies the electrodes included in the ROI. FEP, first-episode psychosis; HC, healthy controls.

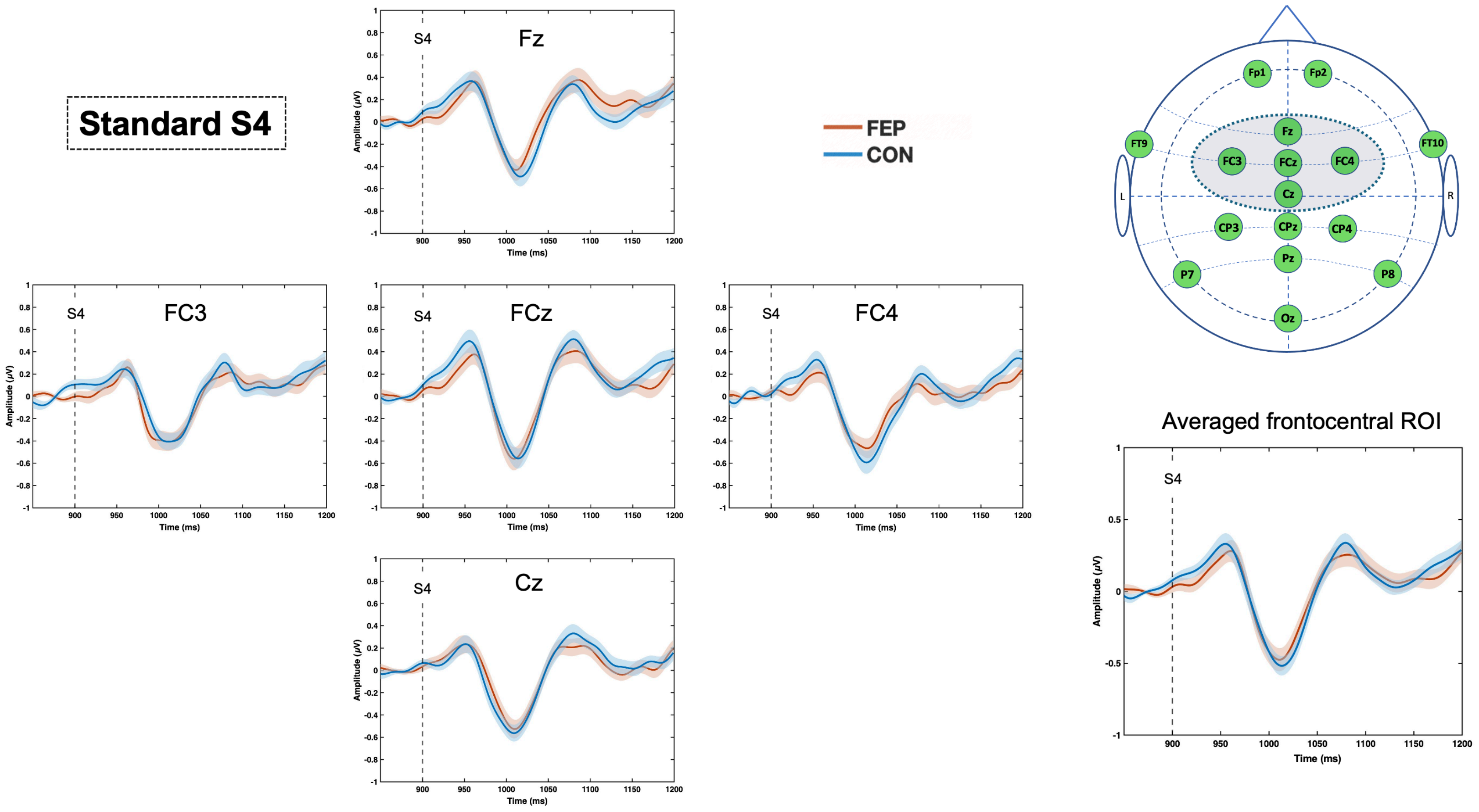

**Figure S4.** Grand-average waveforms elicited by standard S4 tones at five frontocentral electrodes (Fz, FCz, Cz, FC3, and FC4) and across the frontocentral region of interest (ROI). Lines represent group means, and shaded bands indicate ±1 SEM. The vertical dashed line marks S4 onset. The scalp diagram identifies the electrodes included in the ROI. FEP, first-episode psychosis; HC, healthy controls.

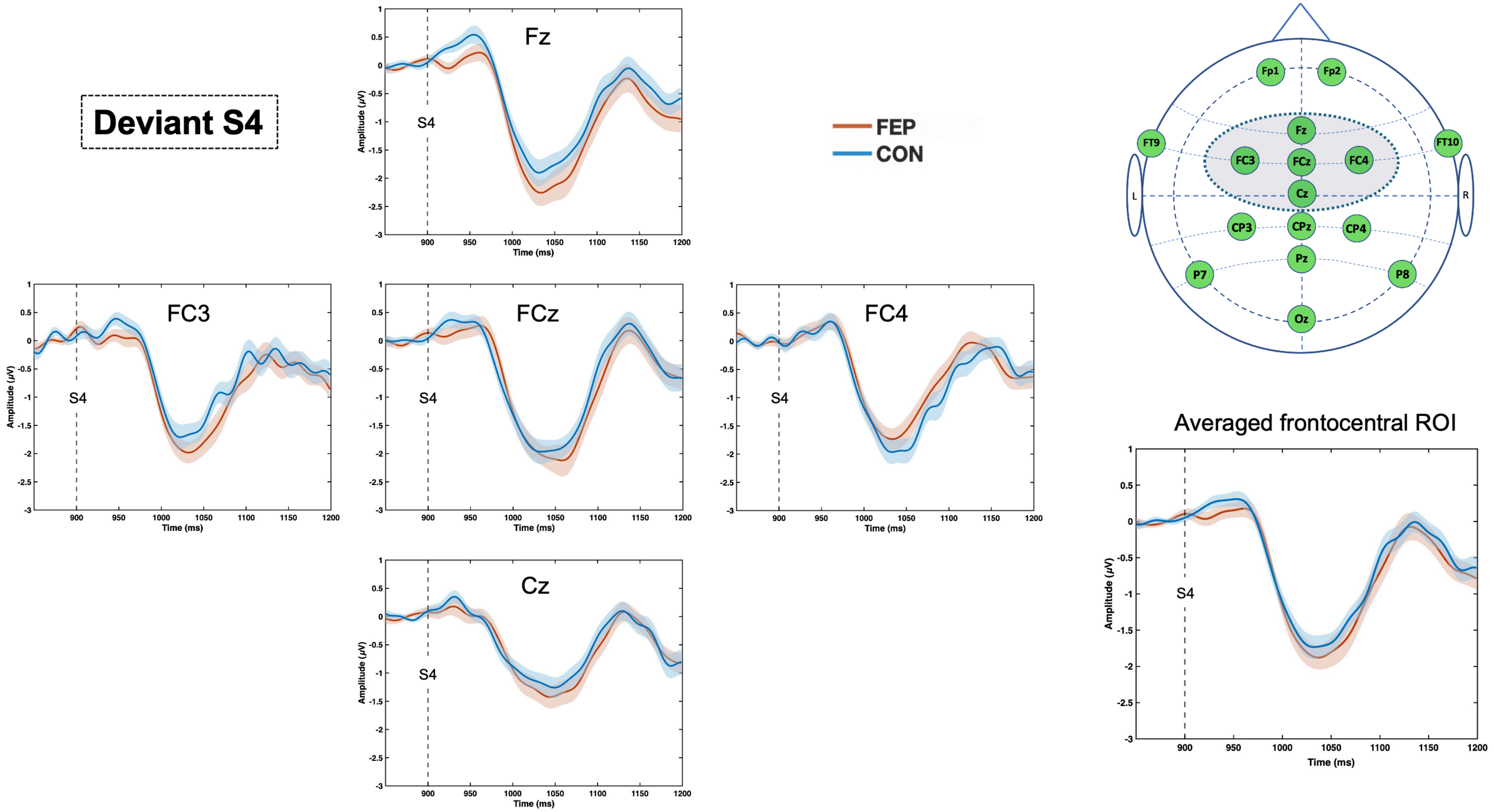

**Figure S5.** Grand-average waveforms elicited by deviant S4 tones at five frontocentral electrodes (Fz, FCz, Cz, FC3, and FC4) and across the frontocentral region of interest (ROI). Lines represent group means, and shaded bands indicate ±1 SEM. The vertical dashed line marks S4 onset. The scalp diagram identifies the electrodes included in the ROI. FEP, first-episode psychosis; HC, healthy controls.
